# Usage of AMH as a determining test of fertility in women with polycystic ovary 2 syndrome (PCOS)

**DOI:** 10.1101/2024.04.29.24306527

**Authors:** Rita Grabowska

## Abstract

PCOS is the most common hormonal disorder and cause of infertility in females of reproductive age. The symptoms and their severity vary strongly between particular cases. PCOS is correlated with hormonal, environmental and genetic factors. Complex interactions between genetics and hormonal levels is important to understand the hormonal 31 abnormality and to assess the chance of pregnancy in women with PCOS.

The research was conducted on patients in the age of 27+/-5 years treated in the 33 Gynecology and Oncology Clinic of CMUJ. The research group - PCOS patients (P) n=62. The control group - (C) n=45. The venous blood was collected in volume of 2 ml centrifuged for 15 min at 1400 rpm. Serum was aspirated to 1.5 ml Eppendorf tubes. The ELISA method was used.

The statistical analysis revealed significant differences in the level of selected factors 38 between the two groups at p <0.01. FSH [IU/ml]: P 5,10 (±1,64) vs K 8,96 (±6,15) LH [ IU/ml]: P 8,59 (±6,79) vs K 11,0 (±6,15) AMH [ng/ml]: P 4,06 (±2,43) vs K 1,47 (±2,14). AMH levels in the PCOS group did not show a significant difference in correlation with age. Obese and overweight women in both 42 groups had significantly different levels of AMH compared with normal-weight women. Furthermore, AMH levels were positively correlated with the age of the first period in the PCOS. The studies indicate a high use of the hormones like FSH and AMH in the diagnosis and assessment of ovarian reserve in women with PCOS.

## Introduction

Polycystic ovary syndrome (PCOS) was discovered in 1844 by Cherau and described by Stein and Leventhal [1]. Despite the intensive research, its pathogenesis remains largely unknown [2]. PCOS occurs in females of all ethnic backgrounds and is the most common hormonal disorder and cause of infertility in females of reproductive age [2,3]. The symptoms include irregular menstrual cycles, hirsutism, acne, and obesity and their 57 severity vary strongly among particular cases [4].PCOS is correlated with hormonal, environmental and genetic factors [5]. Many authors describe complex interactions between genetics and hormonal levels. To assess the chance of pregnancy in women with PCOS it is important to understand the hormonal abnormality which is often done using the antymullerian hormone (AMH) and follicle62 stimulating hormone (FSH) tests [6]. FSH is responsible for the growth and development of the preantral follicles and “protects” them from apoptosis [7,8]. In addition, this hormone activates the aromatase P450, which is responsible for the conversion of the androgens to the estradiol in the cortex of the ovary [9]. The granulosa cells under the influence of FSH begin to produce gonadotrophin surge-inhibiting factor (GnSIF), which by means of the negative feedback begins to inhibit the release of the luteinizing hormone (LH), which plays a key role in the process of ovulation and luteinisation [10,11]. Preantral follicles under the influence of FSH also begin to produce inhibin α (INHα) and inhibin β (INHβ), which primarily inhibit FSH secretion by the gonadotropic cells. LH can stimulate the secretion of inhibins (INHs) by granular-lutein cells after the induction of LH receptors’ expression by FSH [27]. Antimullerian hormone (AMH) is produced exclusively by granulosa cells of ovarian follicles [12] and may have a regulatory effect on the folliculogenesis. Experiments in mice suggest that AMH inhibits the initiation of growth in the primary follicles [13,14], but it is also involved in the growth regulation of antral, 76 preantral and small follicles by inhibiting their sensitivity to FSH [10]. In his experiments, showed that AMH inhibits the epidermal growth factor (EGF), which promotes proliferation of human granulosa cells, which is reflected in the regulation of aromatase P450 activity by 79 the expression of FSH and LH receptors [15,16]. The inhibitory effect of AMH through the acquisition of sensitivity to FSH by the ovary follicles in the later stage of folliculogenesis was confirmed in in vivo studies [15,17]. Moreover, AMH may be involved in regulating the cyclic recruitment of follicles during the transition from the luteal-menstrual phase [18,19]. So far, in women there is sufficient evidence that AMH in the preantral and antral follicles is responsible for its desensitization 85 on FSH [20]. All of these factors are responsible for the proper development of follicles and their growth and functioning. In addition, AMH and FSH increasingly are considered as predictors of ovarian reserve, and when reproductive ovarian function is disturbed these factors can be 89 used as biomarkers of existing pathological conditions.

## The Aim

The aim of this study was to assess the distributions of FSH, LH and AMH levels related to age, the age of the first menarche and the body mass index (BMI) of women with PCOS.

## Subjects and methods

The study was conducted on two groups of patients with an average age of 32+/-5 years treated in the Gynecology and Oncology Clinic of CMUJ, Krakow. The research has been approved by the bioethical commission of UJ CM (KBET/21/B/2009).

Patients were qualified for the study based on their medical history, a general medical 101 examination, a gynecological examination, and their agreement to take part in the research expressed by signing the Patient Aware Consent form. Patients were diagnosed with PCOS by using the Rotterdam criteria listed below (inclusion criteria for group 1).

Blood were taken from PCOS and uterine fibroid patients. These patients had not been treated hormonally during the 6 months preceding the research. BMI was calculated from: body mass/height2

The first group consists of PCOS patients both with and without insulin resistance. The second group is a control group consisting of patients who were not diagnosed with PCOS and were treated by surgery on uterine fibroids (5 laparoscopic myomectomies, 28 myomectomies by laparotomy, 12 total hysterectomy without adnexal laparotomy). Inclusion into the research groups was based on the following criteria:

### Group 1

- irregular periods or secondary amenorrhea
- an enlarged ovary with over 12 small follicles of 2-8mm in diameter (USG-TV ultrasonography-TV examination)
- biochemical symptoms of hyperandrogenism (dehydroepiandrosterone sulfate [DHEAS], testosterone [T], dehydroepiandrosterone [DHEA])

### Group 2 (control group)

- ruled out PCOS
- negative Oral Glucose Tolerance Test (OGTT)

The first research group consisted of PCOS patients (P) n=62. The second group was 125 composed of patients without PCOS and infertility suffering from uterine fibroma (C) n=45. 2 ml of venous blood was collected. Immediately after collection the samples were centrifuged for 15 min at 1400 rpm. Serum was aspirated and transferred to 1.5 ml eppendorf tubes and stored 3 months at -20 C. Baseline blood samples were collected between days 3 and 7 of the menstrual cycle. The biochemical analysis was performed by the ELISA method. The 96-well plates (Biokom) were incubated with the blood serum (50 μl per well) for 12 h at 4 Celsius degrees. Each plate was washed of 3x200 μl in PBS (pH 7.0) using an automatic scrubber. The plates were incubated with the primary antibody at a dilution of 1:10,000 (Thermo Scientific). The time of incubation was 12 h at the temperature of 4 degrees. After that the next step was a 2h incubation with the secondary antibody labeled with HRP (Horseradish peroxidase) and a 15 minute incubation with TMB (3,3′,5,5′137 tetramethylbenzidine, Thermo Scientific). Immediately after adding stop solution (1 nM HCL, Thermo Scientific) the plates were read by using an ELISA reader with the KC Junior program. The theoretical sensitivity of each method was 0.009 ng/ml.

Measurements were performed in duplicate. The standardization curve included 8 steps at the stage r = 0.95.

### Statistical methods

Group values for outcome measures are reported as the mean +/- SD (standard deviation).

Variables which were not normally distributed were log transformed before analysis by parametric methods. Anthropometric data as well as hormonal concentrations were compared by using the Student’s t-test. The relationships between serum AMH, LH, FSH, age and first period as independent variables were assessed by simple and/or multiple linear regression analysis, and Pearson (r) correlation coefficients were presented. Two sided significance levels were reported. Differences were considered significant at the P<0.05. Statistical analyses were performed using STATISTICA software v. 10 (StatSoft, USA, 2011).

## Results

The women of the two groups did not differ significantly in age and BMI (Table 1). FSH levels were significantly lower in women with PCOS compared with control subjects (P < 0.001) Table 1. LH levels were higher in the control group than in the PCOS group (P < 0.001) Table 1. The concentrations of AMH were significantly higher in the PCOS group compared with the control group (P < 0.001) Table 1.

**TABLE 1.**
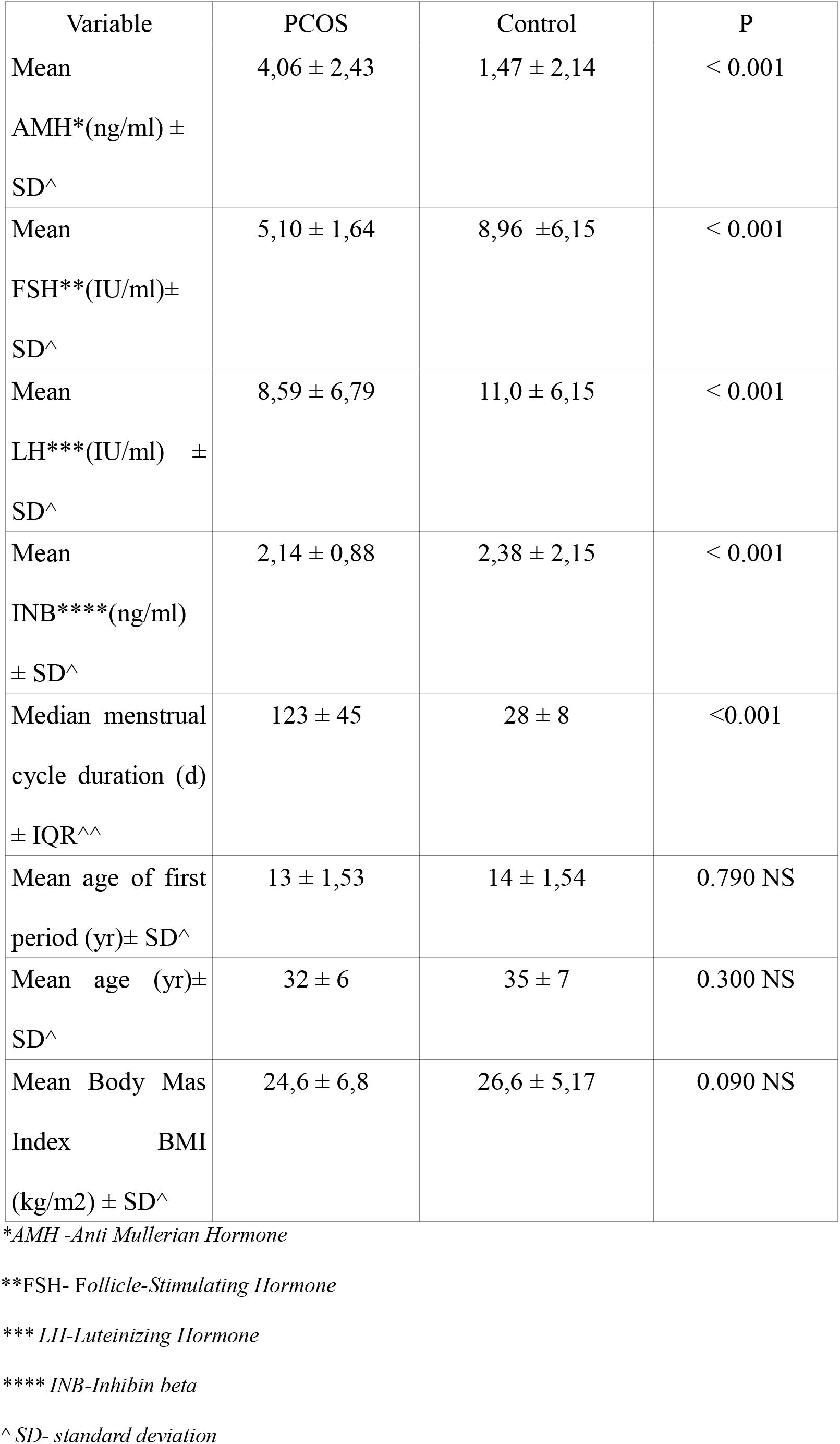
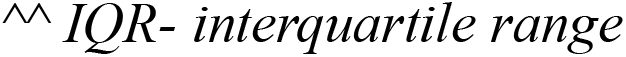
Baseline characteristics and hormone levels of the control group women and women with PCOS.

| Variable | PCOS | Control | P |
| --- | --- | --- | --- |
| Mean<br>AMH*(ng/ml) ±<br>SD^ | 4,06 ± 2,43 | 1,47 ± 2,14 | < 0.001 |
| Mean<br>FSH**(IU/ml)±<br>SD^ | 5,10 ± 1,64 | 8,96 ± 6,15 | < 0.001 |
| Mean<br>LH*** (IU/ml) ±<br>SD^ | 8,59 ± 6,79 | 11,0 ± 6,15 | < 0.001 |
| Mean<br>INB****(ng/ml)<br>± SD^ | 2,14 ± 0,88 | 2,38 ± 2,15 | < 0.001 |
| Median menstrual<br>cycle duration (d)<br>± IQR^^ | 123 ± 45 | 28 ± 8 | <0.001 |
| Mean age of first<br>period (yr)± SD^ | 13 ± 1,53 | 14 ± 1,54 | 0.790 NS |
| Mean age (yr)±<br>SD^ | 32 ± 6 | 35 ± 7 | 0.300 NS |
| Mean Body Mas<br>Index BMI<br>(kg/m2) ± SD^ | 24,6 ± 6,8 | 26,6 ± 5,17 | 0.090 NS |

\*AMH -Anti Mullerian Hormone
\*\*FSH- Follicle-Stimulating Hormone
\*\*\* LH-Luteinizing Hormone
\*\*\*\* INB-Inhibin beta
^ SD- standard deviation
^^ IQR- interquartile range

FSH [ IU/ml] : P 5,10 (±1,64) vs K 8,96 (±6,15) LH [ IU/ml]: P 8,59 (±6,79) vs K 11,0 (±6,15) 162 AMH [ ng/ml]: P 4,06 (±2,43) vs K 1,47 (±2,14) (Table 1).

### AMH determinants

In the control group AMH levels were negatively correlated with age (r = −0,81, P < 0,001) and there was no significant difference in BMI. AMH levels in the PCOS group did not show a significant difference in correlation with age. Obese and overweight women in both groups (Control, PCOS) had significantly different levels of AMH compared with normal weight women (Table 2). We found a significant difference between normal-weight women with PCOS and obese women from the control group in the AMH levels (Table 2). We can find the same situation between over-weight PCOS women and both over-weight and obese control group women. Also significant differences in AMH levels are presented between obese PCOS and obese control group women (Table 2). LH and FSH levels in the two groups did not differ significantly in obese and overweight women compared with normal-weight women.

**TABLE 2.** Body mass effect on the AMH level.

|  | normal-weight | over-weight | obese |
| --- | --- | --- | --- |
| AMH*<br>PCOS**<br>[ng/ml] | 4,35 ± 3,40 A | 4,7 ± 2,4 0<sup>^</sup> aB | 1,31 ± 0,37 <sup>^</sup> b |
| AMH*<br>control<br>[ng/ml] | 2,33 ± 2,00 | 1,21 ± 0,32 ^^ a | 0,74 ± 0,13 ^^ ABb |

\*AMH -Anti Mullerian Hormone
\*\*PCOS-Polycystic Ovary Syndrome
<sup>^</sup> - Significantly difference at $P < 0.01$
^^ Significantly difference at $P < 0.01$
A- Significant difference at $P < 0.05$
a -Significant difference $P < 0.01$
B – Significant difference at $P < 0.01$
b- Signficant difference at $P < 0.01$

### Interactions were not observed

Furthermore, AMH levels were positively correlated with the age of the first period in the PCOS group (r = 0,43, P <0,01) but there was no significant correlation in the control 181 group. The number of periods in the PCOS group was not correlated with levels of AMH but in the control group a negative correlation was shown between the number of periods and AMH levels (r = - 0,71, P < 0,01). We presented a positive correlation between LH levels and AMH levels in the PCOS group, r = 0,6282; P < 0,001 and no correlation between LH 186 levels and AMH levels in the control group.

## Discussion

The study showed that AMH serum levels were significantly higher in PCOS women compared with non PCOS women – the same results can be found at Joop et al. and Arabzadeh’s et al. [21,22]. The high levels of AMH in the PCOS group can be connected with an ovarian dysfunction. It is well-known that the ovulation problems in PCO syndrome is connected with an increase in early follicular growth and poor selection of Graff follicle from the increased pool [24]. A large number of immature follicles might be a source of increased production of AMH [23]. It has been shown that granulosa cells (GC) from polycystic ovaries produced 75 times more AMH than GC from healthy ovaries so it suggests that not only excessive accumulation of the antral follice can have effect on AMH level [12, 29]. LH levels were lower in women with PCOS than in women from the control group. Therefore, we showed that the FSH level is significantly lower in the PCOS group compared to the control group in contrast to Codne et al. but we had the same results as 200 Piouka et.al. [24, 26]. The lower level of FSH and LH in PCOS women can be an effect of AMH high level which plays a important role as an inhibitory factor of steroidogenesis causing lower E2 values and it is well known that FSH and LH level is regulated by negative feedback action of E2 hormone [21, 28].

With every menstrual cycle AMH level should decrease but our study showed that there was is no significant correlation between AMH levels and the number of periods and also no correlation between AMH levels and age in the PCOS group. It can be the result of a higher primordial follicle pool at birth time or lower diminishing levels of follicle pool during childhood and menarche – Pascal et al. in his data also reported no relationship between AMH levels and age in the PCOS. Results of our study showed the relationship between AMH levels, age and the number of periods in the control group and data of Pascal and Woo et al. showed the same results between age and AMH levels [30]. We found the significant correlation between AMH levels and the age of the first period – it is novel idea to think the role of studyin AMH. The hypothesis of that is: women with PCOS can keep their ovary reproductive function longer than healthy women because of a constant level of AMH during reproductive age. The date of the study did not show the relationship between AMH levels and the first period in the control group. Our observation showed that AMH concentration was not significantly influenced by LH levels in the control group but was significantly correlated in the PCOS group which were the same results obtain by Rooji et all [21]. We did not find a relationship between FSH and AMH levels in the PCOS group but a positive correlation was observed in the control group in contrast to Pascal et al. and 221 Neoklis et al. [15, 16].

There was also no correlation between LH, AMH and FSH levels between obese, over223 weight and normal-weight women in the PCOS group in contrast to the research data of Pascal in which is showed the significant differences between obese and non-obese women with PCOS [15]. Observation of Piouka et al. agree with our study [26]. Although, no significant association between AMH levels and BMI was observed in the PCOS group.

In conclusion, AMH levels are significantly increased in PCOS which makes it a good marker for this syndrome. Moreover, high AMH level maintains ovary reproductive function in a good condition longer in PCOS women than in healthy women. Increased AMH levels in women with PCOS seem to be merely the result of increased small follicle number and directly increasing number of GC. We can say that AMH is the most important independent marker which determines the ovary function. It should be emphasized, though, that a causal relationship is at present difficult to establish and more studies are definitely needed.

## Data Availability

All data produced in the present work are contained in the manuscript

## Acknowledgments

I would like to thank the Dean of Medicine Faculty, Jagiellonian University Collegium Medicum, for financial support of this study (K/ZDS/000070).

